# Apramycin resistance in bacteria isolated from humans, a systematic review and meta-analysis

**DOI:** 10.1101/2025.10.20.25338358

**Authors:** J Scott Weese, Heather E Weese

**Author notes:** Address correspondence to J Scott Weese.

## Abstract

**Introduction:** Apramycin is an aminoglycoside antimicrobial that has been used in veterinary medicine since the 1980s but not licensed for human medicine. Because it is not impacted by common aminoglycoside resistance mechanisms, there is interest in repurposing the drug for use in humans, as a treatment of multidrug resistant Gram negative bacterial infections.

**Gap Statement:** The prevalence and factors associated with apramycin resistance in bacteria isolated from humans has received limited study but is important foundational information for considering repurposing apramycin for use in humans.

**Aim:** To systematically review and analyze data pertaining to apramycin resistance in bacteria isolated from humans and to identify knowledge gaps, to inform work evaluating the potential for re-purposing of apramycin for clinical use in humans.

**Methodology:** A systematic review was performed to evaluate apramycin resistance in bacteria isolated from humans.

**Results:** A total of 1626 references were identified during the search, with 34 studies were deemed eligible for inclusion. Pooled estimates for apramycin resistance were 6% (95% CI 1-12%) for *E. coli,* 1% (0-3%) for *Acinetobacter* spp, 2% (0-5%) for *Enterobacter* spp, 7% (2-15%) for *Klebsiella* spp, 4% (0-13%) for *Pseudomonas* and 0% (0-0%) for *Salmonella* spp. Multivariable mixed-effects meta-regression identified no effect of year (*P*=0.36), bacterial species (all *P*>0.19), geographic region (all *P*>0.13) or enrollment of known carbapenem-resistant isolates (*P*=0.44). The only significant variable was datasets that used known gentamicin-resistant isolates (*P*=0.003).

**Conclusion:** Despite nearly 50 years of apramycin use in animals, apramycin resistance was identified in bacteria of human origin but was rare, supporting the potential value of re-purposing this drug for use in humans and suggesting that there is limited spillover of resistance from veterinary and agricultural use of apramycin.

---

The silent pandemic of antimicrobial resistance (AMR) poses increasing challenges as multidrug resistant bacterial infections increase and millions of lives are impacted annually.^1^ A multimodal approach is required to address AMR, and part of that involves development of new treatment options. Discovery and testing of new antimicrobial molecules are slow and very expensive, and this process can potentially by repurposing drugs that were originally developed and abandoned or were only pursed in the veterinary market. One example of this is apramycin, a veterinary aminoglycoside that has emerged as a potentially appealing target for repurposing.

Apramycin was discovered in 1967^2^ and while it has been authorized for use in animals since the 1980s, it was never authorized in humans. As with other aminoglycosides, apramycin has excellent activity against aerobic Gram negative bacteria, and, also like other aminoglycosides, is potential ototoxic and nephrotoxic. These concerns, along with development of other drug classes with excellent activity against Gram negative pathogens during that era, likely explain the lack of authorization of the drug in humans at that time. However, as AMR increases in Gram negative pathogens, there has been renewed interest in apramycin in humans. This is in large part because the unique structure of apramycin, compared to currently used aminoglycosides, enables it to resist inactivation by the common aminoglycoside-modifying enzymes, even those typically found in multidrug-resistant (and extensively drug-resistant) Gram-negative bacteria.^3^

Apramycin is currently used in animals in many countries, for treatment or prevention of enteric infections in livestock and, to a limited degree, for non-veterinary use (growth promotion). The role of antimicrobial use in animals in development of resistance in human pathogens is poorly understood but there are clear examples where veterinary or agricultural use has contributed to resistance in human pathogens.^4,5^ This raises the question of whether veterinary and human use of apramycin could co-exist without veterinary use creating unacceptable risk of resistance emergence in human pathogens. Such an assessment is complicated and requires an understanding of the use of the drug in animals, the importance of the drug in animals, antimicrobial resistance in bacteria from animals and antimicrobial resistance in bacteria from humans. The objective of this study was to systematically review and analyze data pertaining to apramycin resistance in bacteria isolated from humans and to identify knowledge gaps, to inform work evaluating the potential for re-purposing of apramycin for clinical use in humans.

## Materials and Methods

A systematic review was designed based on Preferred Reporting Items for Systematic Review and Meta-Analysis Protocols (PRISMA-P) guidelines.^6^ The full protocol is available at the University of Guelph’s Atrium Institutional Repository (https://hdl.handle.net/10214/29094).

The review aimed to address the following CoCoPop (condition, context, population) and CoCoPop-T (condition, context, population, time) questions:

*CoCoPop1: What is the prevalence of apramycin resistance in bacteria isolated from humans?*
*CoCoPop2: Are there differences in the prevalence of apramycin resistance in different bacterial species isolated from humans?*
*CoCoPop-T: Has the prevalence of apramycin resistance in bacteria isolated from humans changed over time?*

### Search strategy

CAB Abstract, Medline (via OVID) and Web of Science were searched. A broad search strategy was used, querying “apramycin” in all fields. Reference lists of identified reviews and included manuscripts were manually searched to identify additional relevant citations. The search was initially performed July 4, 2024 and updated June 25, 2025.

### Reference screening

Search results were imported into Covidence (Covidence systematic review software, Veritas Health Innovation, Melbourne, Australia. Available at www.covidence.org) and de-duplicated. Title and abstract screening were performed by two reviewers using the following criteria.

### Title and abstract screening

1. Does the study describe the prevalence of apramycin resistance in bacterial isolates from humans (excluding studies evaluating known apramycin-resistant isolates?)
2. Is it a prospective observational study, controlled trial or other primary research study? Case reports, case series’, basic microbiological or mechanistic studies and pharmacokinetic studies were excluded.

Reviewers discussed any conflicts and came to consensus, being blinded as to their original assessment. A calibration step was performed after approximately 10% of the title/abstract search has been performed by both reviewers, with a plan to review the evaluation criteria if there was <90% agreement.

Full text screening was then performed by two reviewers, using the following review criteria.

### Full text review

1) Is the full text available?
2) Is it a prospective observational study, randomized controlled trial or other primary research study?
3) Does it describe the prevalence or incidence of apramycin resistance in bacterial isolates from healthy and/or diseased humans? Conflicts were reviewed and resolved via discussion.

### Data extraction

Data extraction was performed by two reviewers into a pre-tested spreadsheet. Information about the study (e.g. authors, year, country), study population (e.g. health status, setting), bacterial species and resistance rates (numerator and denominator) was retrieved. For reporting of year, if yearly data were pooled, the median year from the date range was used if it encompassed 7 years or less. If the range was greater than 7 years, the study was excluded from temporal analysis. Determination of results as susceptible, intermediate or resistant was based on the criteria used by the individual study. Data extraction results were compared, and conflicts were resolved through discussion.

### Critical appraisal of individual sources of evidence

Two reviewers performed risk of bias assessment using the JBI Critical Appraisal Checklist for Prevalence Studies^7^ to determine if studies should be included in analysis. Consensus was used to resolve any disagreements.

### Data analysis

Study characteristics were described. Prevalence meta-analyses were conducted using a random-effects model implemented via the metaprop() function from the **meta** package in R. To stabilize variance across studies, proportions were transformed using the Freeman–Tukey double arcsine method. The inverse variance method was used for pooling, and between-study heterogeneity was estimated using the DerSimonian–Laird estimator. Hartung–Knapp adjustment was applied to improve the accuracy of confidence intervals because of anticipated high heterogeneity. Forest plots were generated to display pooled prevalence with 95% confidence intervals, both overall and within relevant subgroups. Between-study heterogeneity was assessed using the I² statistic and τ². Where substantial heterogeneity was observed, subgroup analyses were conducted to explore potential sources.

Mixed-effects meta-regression was conducted to assess whether bacterial species were associated with the prevalence of antibiotic resistance. Analyses were restricted to bacterial species represented in at least three studies. Meta-regression was performed using the metareg() function from the meta package in R, with bacterial species included as a categorical covariate. Proportions were transformed using the Freeman–Tukey double arcsine method to stabilize variance. Between-study heterogeneity was modeled using a restricted maximum likelihood (REML) estimator. The model estimated the proportion of heterogeneity explained by covariates (R²) and tested moderator significance using F-tests. Residual heterogeneity was evaluated using τ², I², and the Q-test for residual heterogeneity. Statistical significance of individual coefficients was assessed using t-tests.

Multivariable mixed-effects meta-regression was also performed to examine the association between antibiotic resistance prevalence and multiple variables, including year of study, bacterial species, and whether isolate collections were selected based on carbapenem or gentamicin resistance. A *P*<0.05 was considered significant for all analyses.

## Results

A total of 1626 references were identified during the search, with two others identified through review of reference lists. After de-duplication and screening, 34 studies were deemed eligible for inclusion (Figure 1, Table 1). Apramycin resistance rates in more than one bacterium were reported in some studies, so the 34 studies reported data from a total of 61 different bacterium-level analyses.

**Figure 1:**
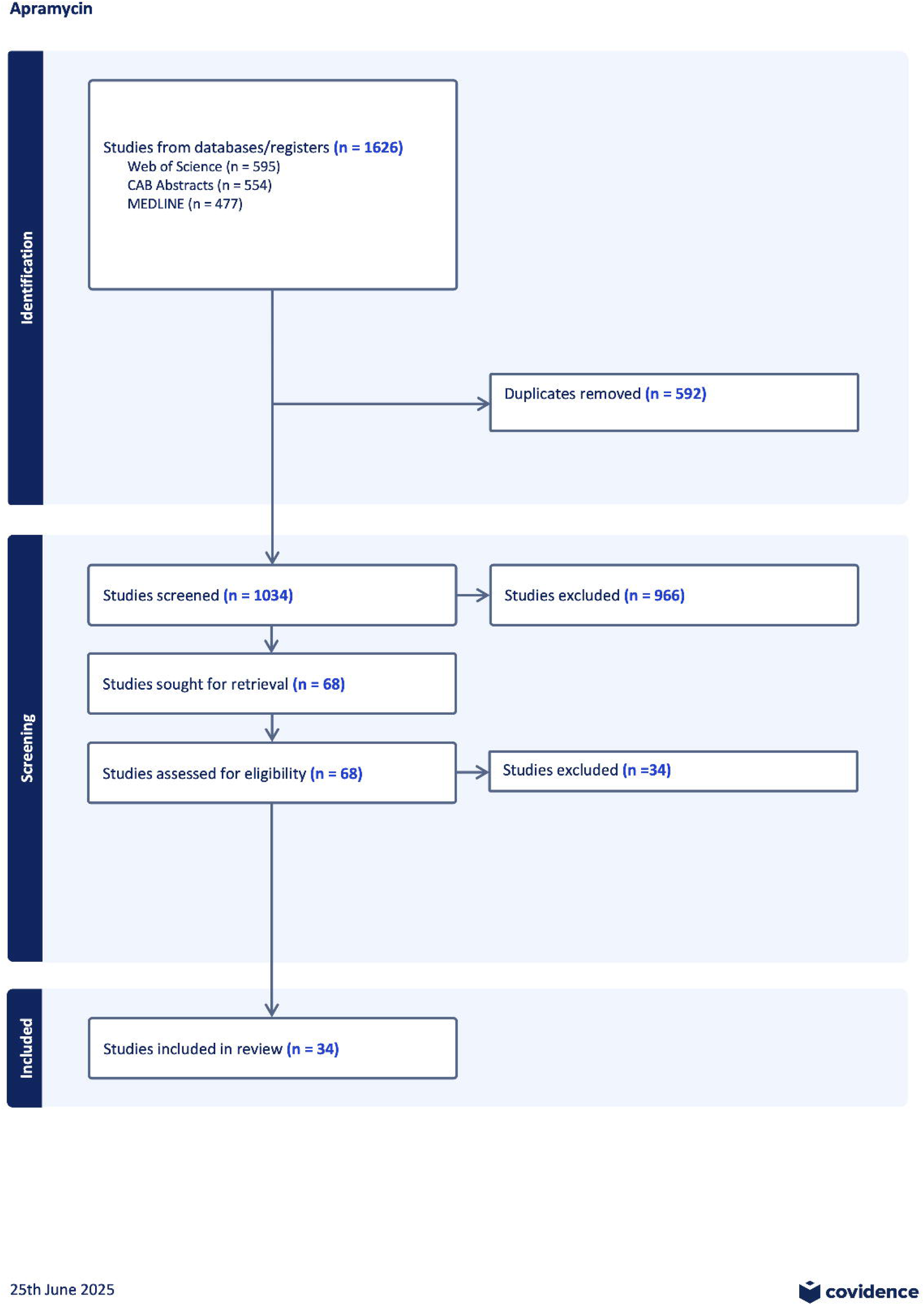
PRISMA diagram showing screening process and final number of studies from which data were extracted for a systematic review of apramycin resistance in bacteria isolated from humans.

**Table 1:**
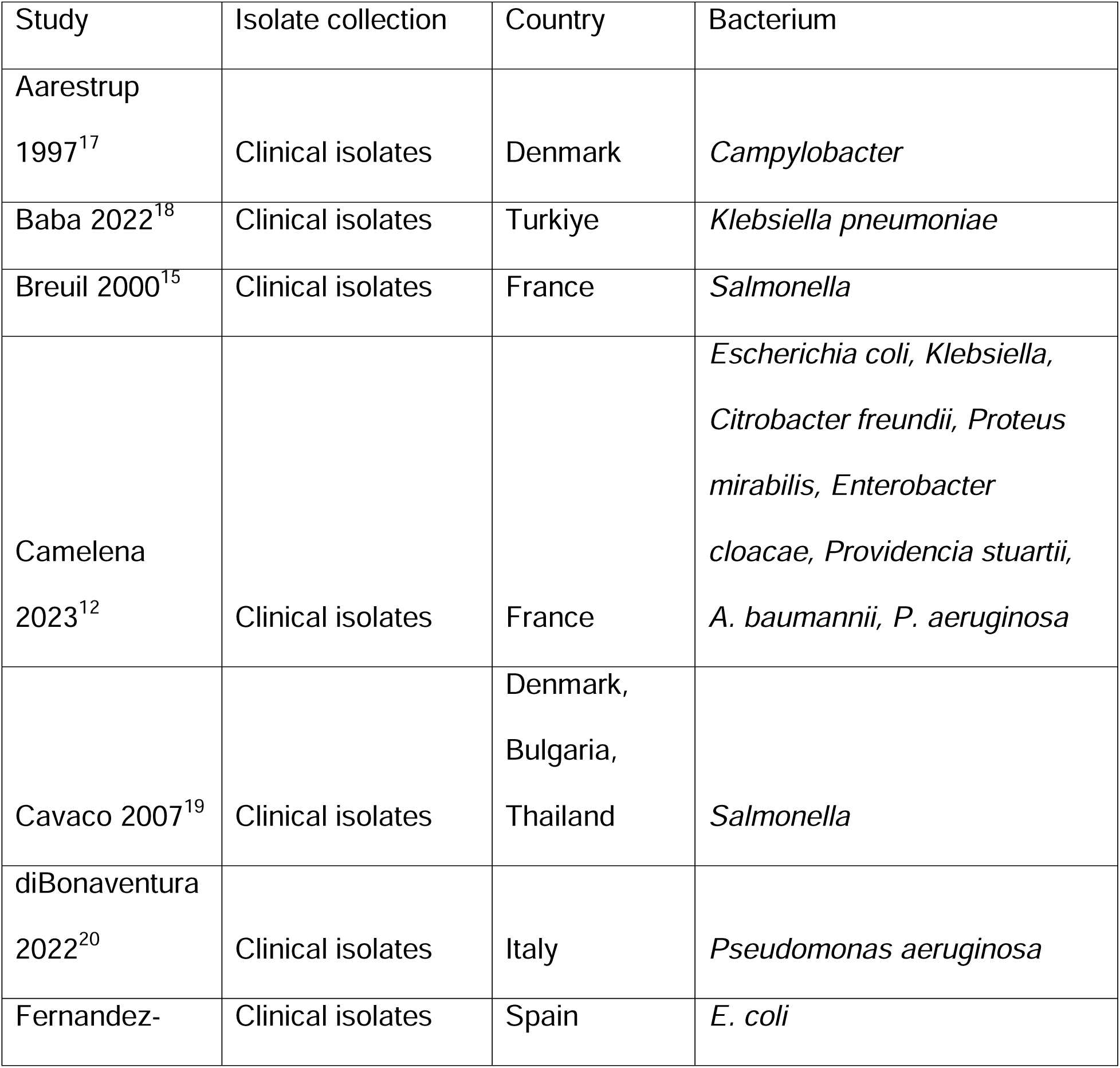
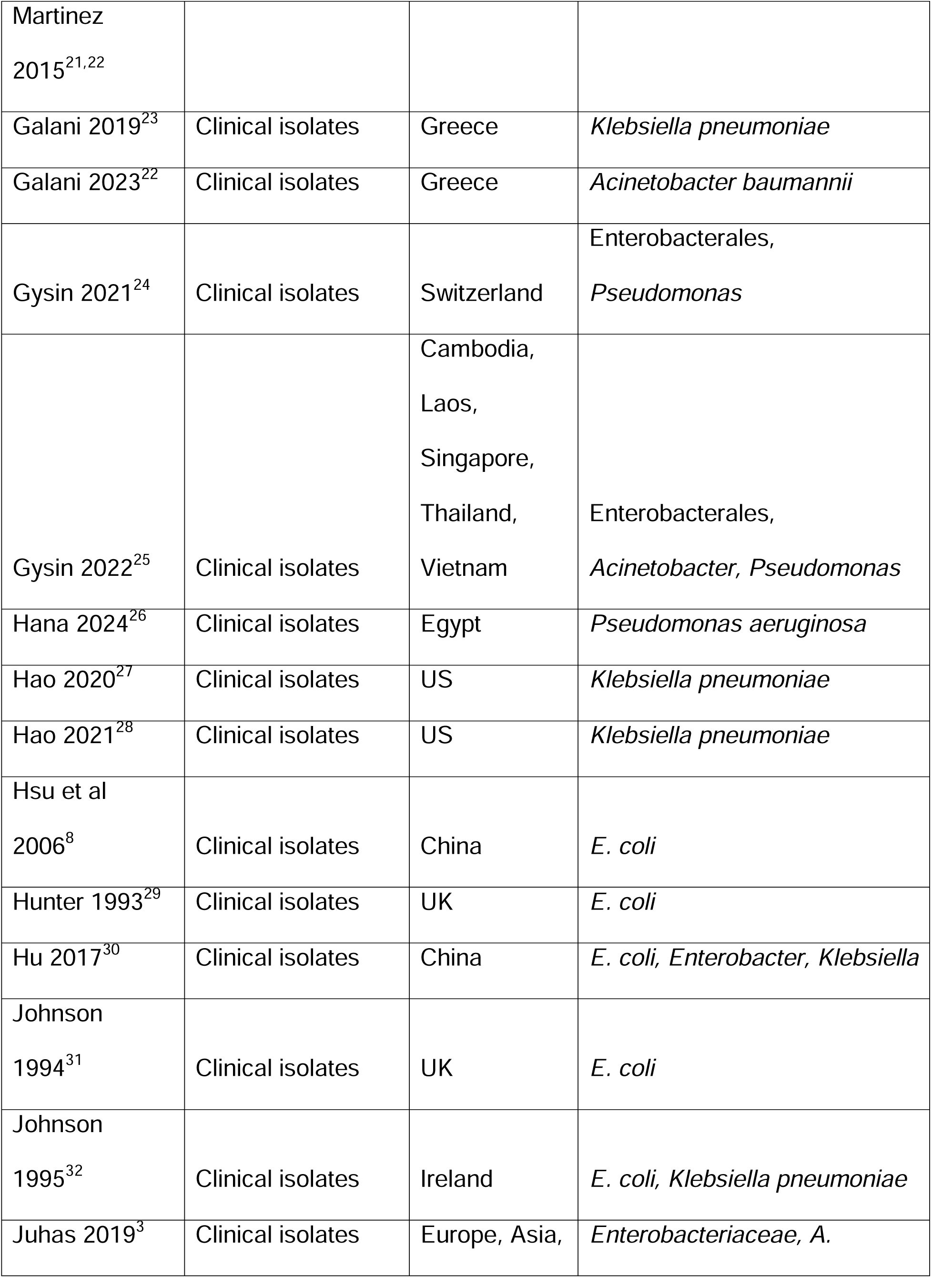
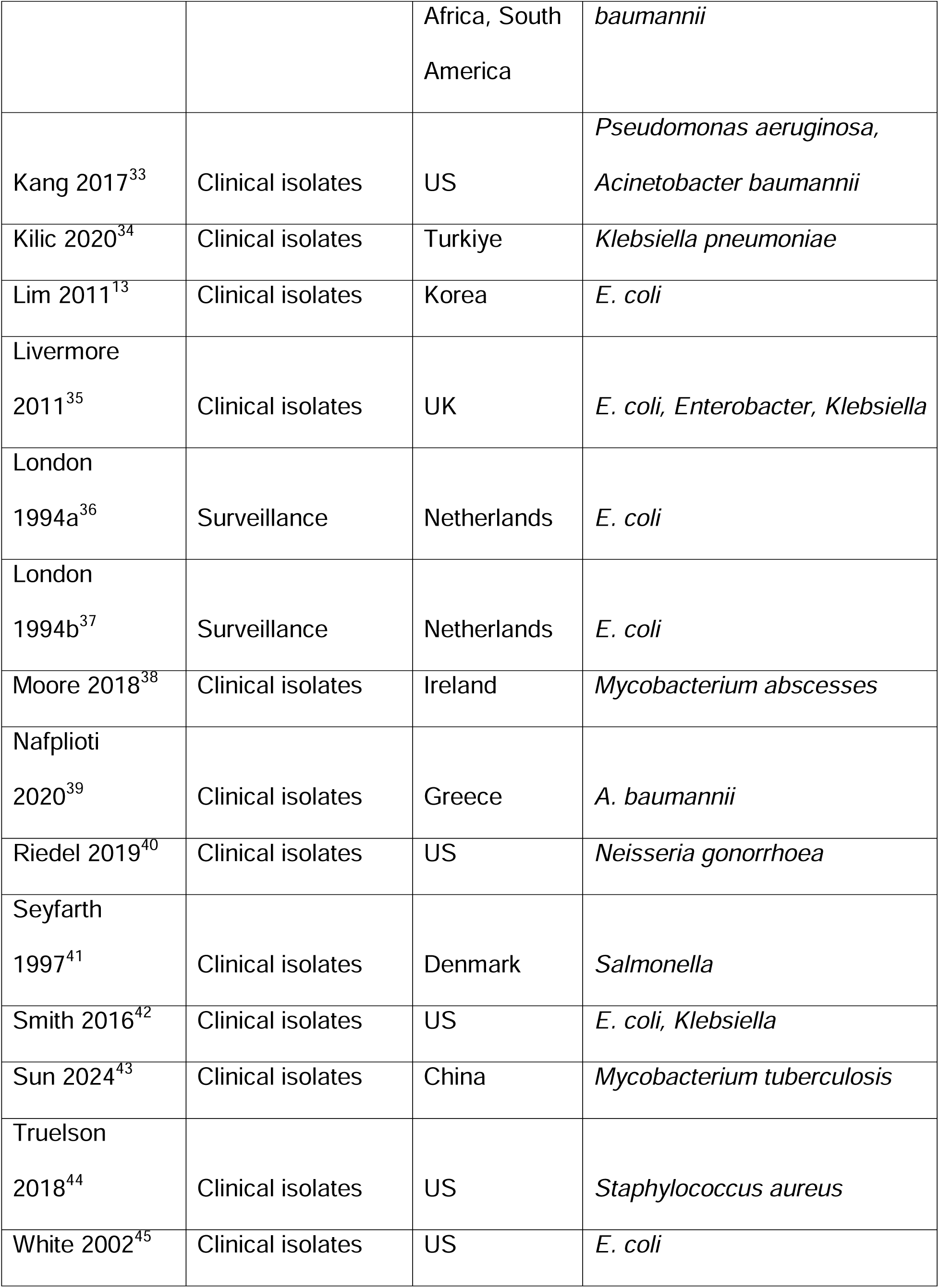
Included studies in a meta-analysis of apramycin resistance in bacteria isolated from humans.

After review of the JBI Critical Appraisal Checklist, all were deemed eligible for analysis. However, many studies contained isolate collections that were biased towards antimicrobial resistance, including 27 populations that consisted of isolates that were all known to be carbapenem-resistant and 16 that were known to be gentamicin resistant. These were included in the overall pool prevalence estimation, but sub-analyses were done, as numbers permitted, for these collections and those not specifically selected as being resistant. Some other isolate collections would be expected to have some bias toward resistance (e.g. isolates from patients with cystic fibrosis, isolates from multidrug resistant bacterium collections) but these were not included in the resistant subgroup because they were not enrolled specifically because of known resistance.

Studies were published between 1993 and 2024 (Figure 2). The 61 pathogen-level analyses involved *E. coli* (n=18, 30%), *Klebsiella pneumoniae* (12, 20%), *Pseudomonas* spp (6, 9.8%), *Acinetobacter* sp. (6, 9.8%) *Salmonella* (3, 4.9%), *Enterobacter* spp (3, 4.9%), unspecified Enterobacterales (3, 4.9%), and one (1.6%) each of *Campylobacter, Citrobacter freundii, Morganella morganii, Mycobacterium abscessus, Mycobacterium tuberculosis, Neisseria gonorrhoea, Proteus mirabilis, Providencia* spp, *Serratia marcescens* and *Staphylococcus aureus*.

*CoCoPop1: What is the prevalence of apramycin resistance in bacteria isolated from humans?*

All identified studies addressed this question. However, there were variable bacterial species and isolate collections (e.g. isolates known to be resistant to specific antimicrobials vs broader prevalence studies.) Pooled prevalence estimates and 95% confidence intervals for all bacteria are presented Table 2. Further analysis was performed only with bacterial species represented by 3 or more studies.

**Figure 2:**
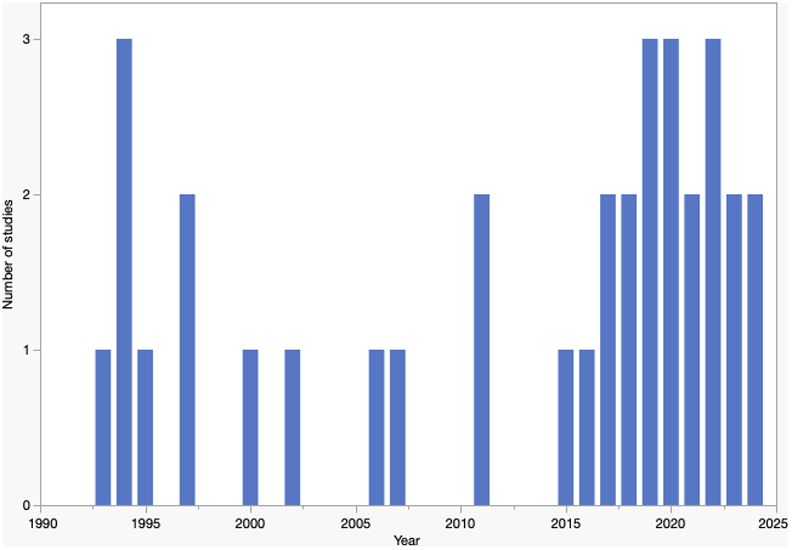
Distribution of publication dates of manuscripts identified in a systematic review of apramycin resistance in bacteria isolated from humans (n=31).

**Table 2:** Pooled prevalence and 95% confidence intervals for the prevalence of apramycin resistance in bacteria isolated from humans (n=60* populations from 34 studies)

| Bacterium (number of studied populations) | Pooled prevalence: proportion (95% confidence interval) |  |  |
| --- | --- | --- | --- |
|  | Overall | Studies not based on selection of carbapenem- or gentamicin resistant isolates (n) | Study of carbapenem- or gentamicin-resistant isolates (n) |
| *Acinetobacter* (n=6) | 0.01 (0.00-0.03) | 0.00 (0.00-0.06)<br>(n=3) | 0.02 (0.00-0.11)<br>(n=3) |
| *Campylobacter* (n=1) | 0.00 (0.00-0.04) | 0.00 (0.00-0.04)<br>(n=1) |  |
| *Citrobacter freundii* (n=1) | 0.02 (0.00-0.06) |  | 0.02 (0.00-0.06)<br>(n=1) |
| *E. coli* (n=17*) | 0.06 (0.01-0.12) | 0.01 (0.00-0.01)<br>(n=8) | 0.14 (0.05-0.27)<br>(n=9) |
| *Enterobacter* (n=3) | 0.03 (0.00-0.17) |  | 0.03 (0.00-0.17)<br>(n=3) |
| Enterobacterales, unspecified (n=3) | 0.01 (0.00-0.07) | 0.00 (0.00-0.02)<br>(n=2) | 0.04 (0.01-0.10)<br>n=1) |
| *Klebsiella* (n=12) | 0.07 (0.02-0.15) | 0.05 (0.00-1.0)<br>(n=2) | 0.07 (0.01-0.17)<br>(n=10) |
| *Morganella morganii* (n=1) | 0.03 (0.01-0.15) |  | 0.03 (0.01-0.15)<br>(n=1) |
| *Mycobacterium abscessus* (n=1) | 0.00 (0.00-0.21) | 0.00 (0.00-0.21)<br>(n=1) |  |
| *Mycobacterium tuberculosis* (n=1) | 0.25 (0.17-0.35) | 0.25 (0.17-0.35)<br>(n=1) |  |
| *Neisseria gonorrhea* (n=1) | 0.00 (0.00-0.05) | 0.00 (0.00-0.05)<br>(n=1) |  |
| *Proteus mirabilis* (n=1) | 0.00 (0.00-0.10) |  | 0.00 (0.00-0.10)<br>(n=1) |
| *Providencia* (n=1) | 0.04 (0.00-0.10) |  | 0.04 (0.00-0.10)<br>(n=1) |
| *Pseudomonas* (n=6) | 0.04 (0.00-0.13) | 0.03 (0.00-0.14)<br>(n=5) | 0.11 (0.02-0.29)<br>(n=1) |
| *Salmonella* (n=3) | 0.00 (0.00-0.00) | 0.00 (0.00-0.00)<br>(n=3) |  |
| *Serratia marcescens* (n=1) | 0.08 (0.02-0.19) |  | 0.08 (0.02-0.19)<br>(n=1) |
| *Staphylococcus aureus* (n=1) | 0.00 (0.00-0.00) | 0.00 (0.00-0.00)<br>(n=1) |  |

\* Excluding one outlier<sup>8</sup> that was removed from analysis

Pooled resistance estimates, with subgroup estimates for isolate populations that did or did not include known carbapenem or gentamicin-resistant isolates were performed. Results for *E. coli* are presented in Figure 3. Very high heterogeneity was present. One study^8^ was a particular outlier, reporting apramycin resistance in 102/110 (93%) isolates. In that study, it was stated that isolates were randomly selected from pre-treatment patient samples, but with the high prevalence of resistance and apramycin resistance almost double the gentamicin resistance rate (47.3%), there were concerns about the validity of the results. Removal of that study resulted in a drop of heterogeneity of the subpopulation, as measured by I^2^, from 98.6% to 0%, and a decrease in the pooled prevalence from 7% (95% CI 0-34%) to 1% (0-2%) (Figure 5). This study was removed from further analysis.

**Figure 3:**
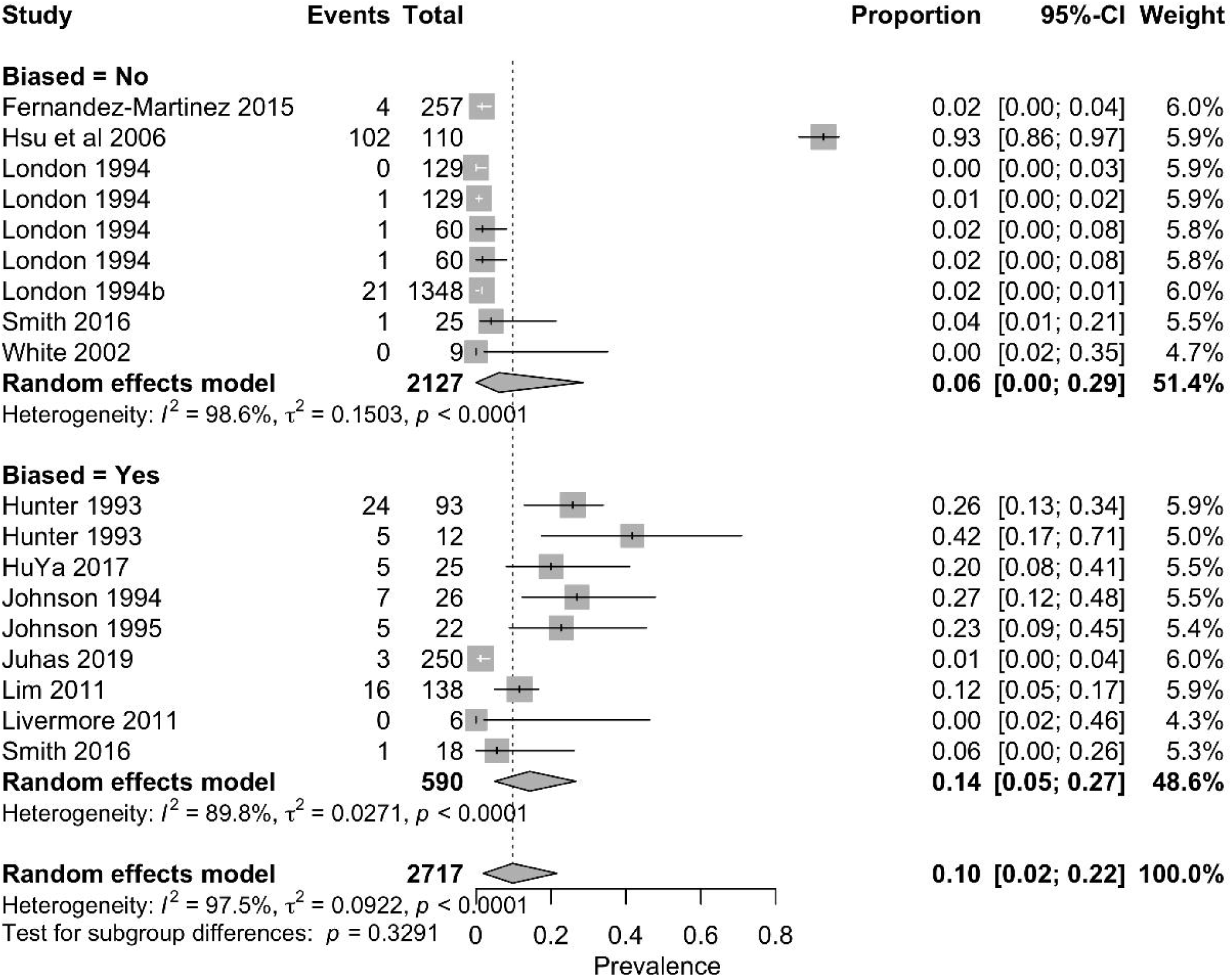
Meta-analysis presenting the pooled prevalence and 95% confidence intervals for apramycin resistance in *Escherichia coli* from, a) 18 isolate collections that enrolled known multidrug resistant isolates (n=9) and those that did not (n=9)

**Figure 4:**
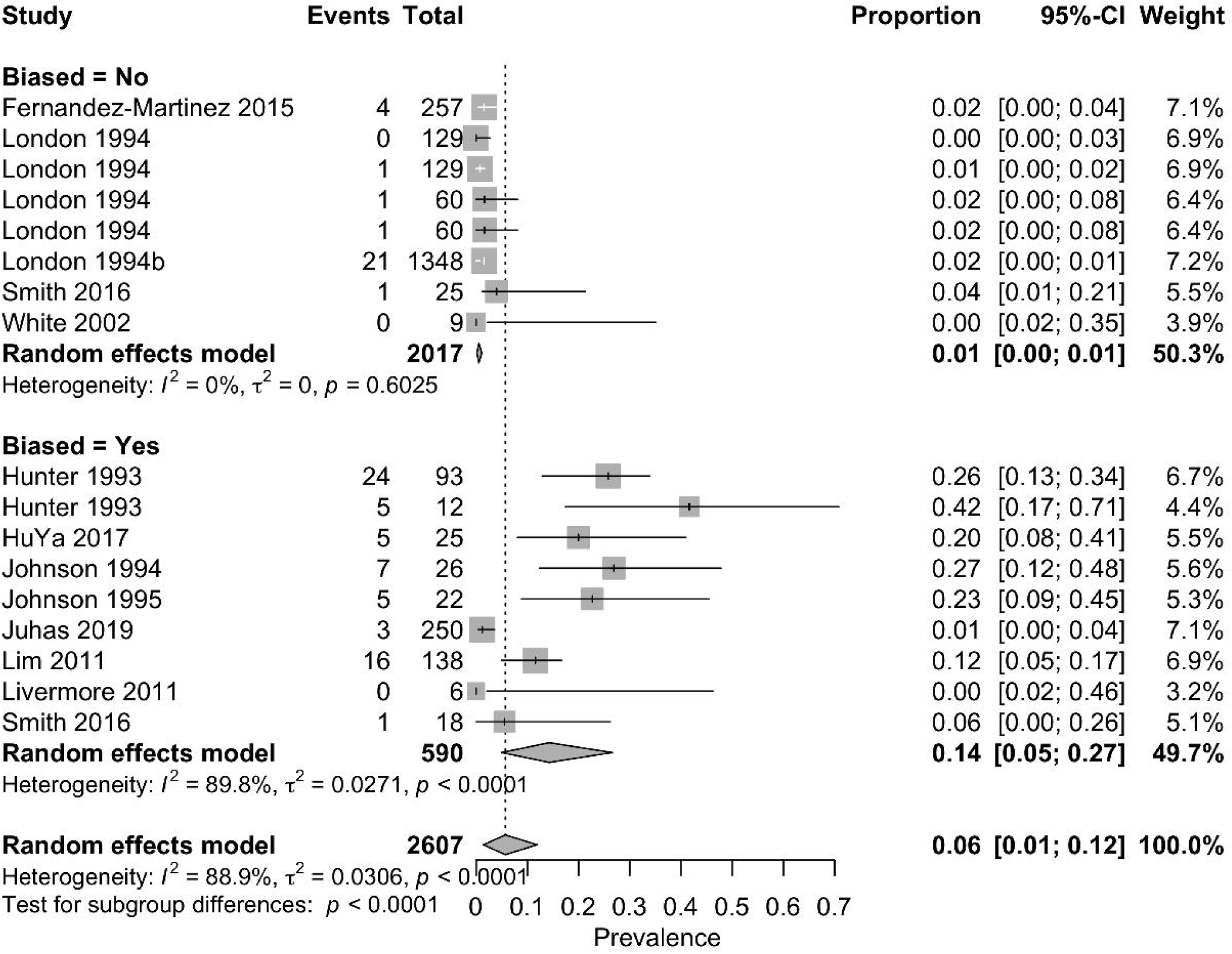
Meta-analysis presenting the pooled prevalence and 95% confidence intervals for apramycin resistance in *Escherichia coli* from 17 isolate collections after removal of an outlier

Pooled estimate and 95% confidence intervals for *Acinetobacter, Enterobacter, Klebsiella, Pseudomonas* and *Salmonella* are presented in Figures 5-9, respectively. Studies of *Klebsiella* focused largely on resistant isolate collections, with 10/12 (83%) being carbapenem resistant and/or gentamicin resistant isolates. All studies of *Enterobacter* spp were of carbapenem resistant collections. In contrast, the pooled estimate for *Salmonella* was based on 3 studies, none of which were selected for resistant isolates, with no apparent heterogeneity. When collections that were selected as gentamicin or carbapenem-resistant are removed, the pooled prevalence for bacteria that were represented by at least 3 studies was 3% (95% CI 0-10%) (Figure 10).

**Figure 5:**
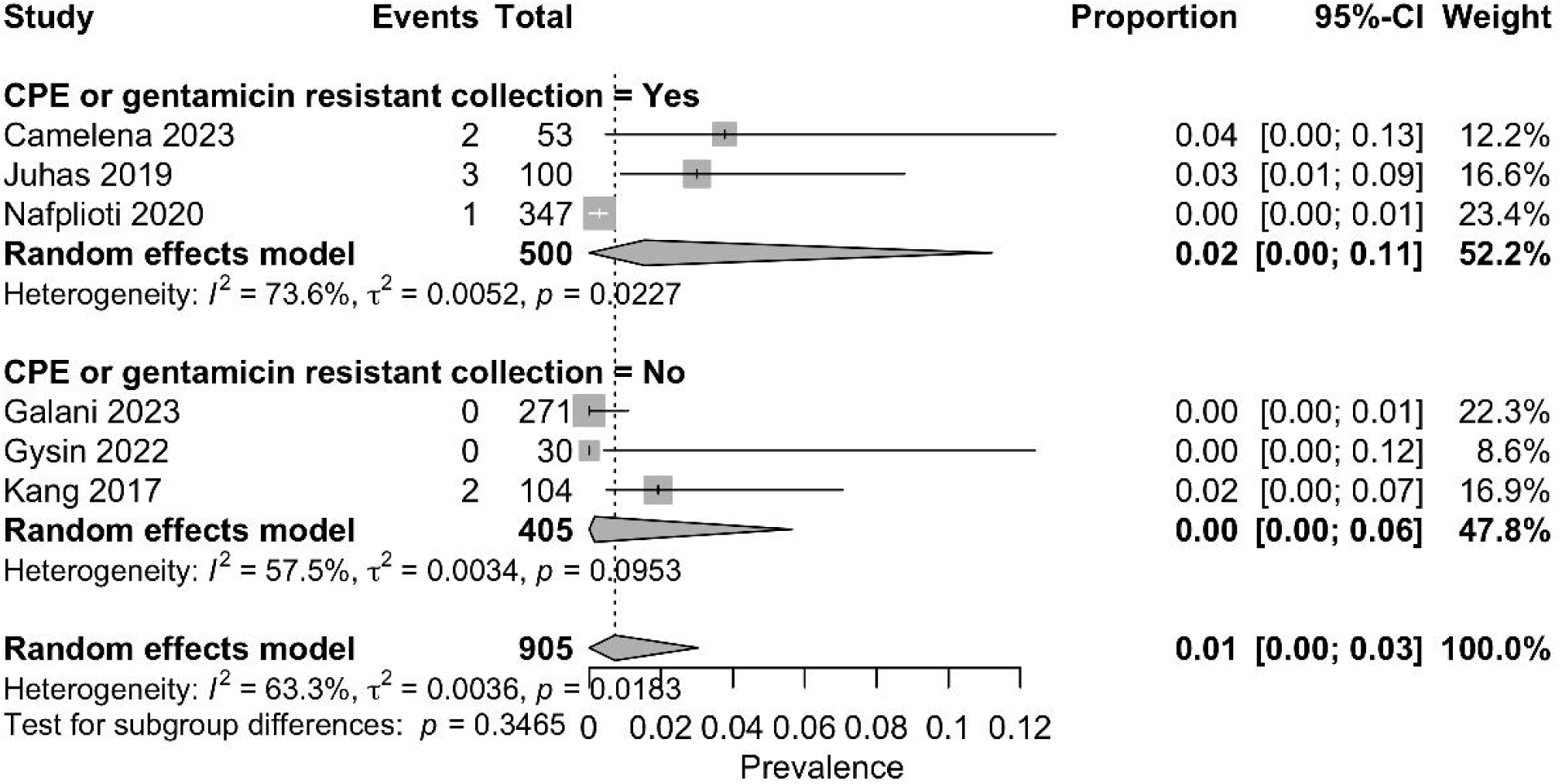
Meta-analysis presenting the pooled prevalence and 95% confidence intervals for apramycin resistance in *Acinetobacter* spp (n=6 isolate collections) from studies that enrolled known multidrug resistant isolates (n=3) and those that did not (n=3)

**Figure 6:**
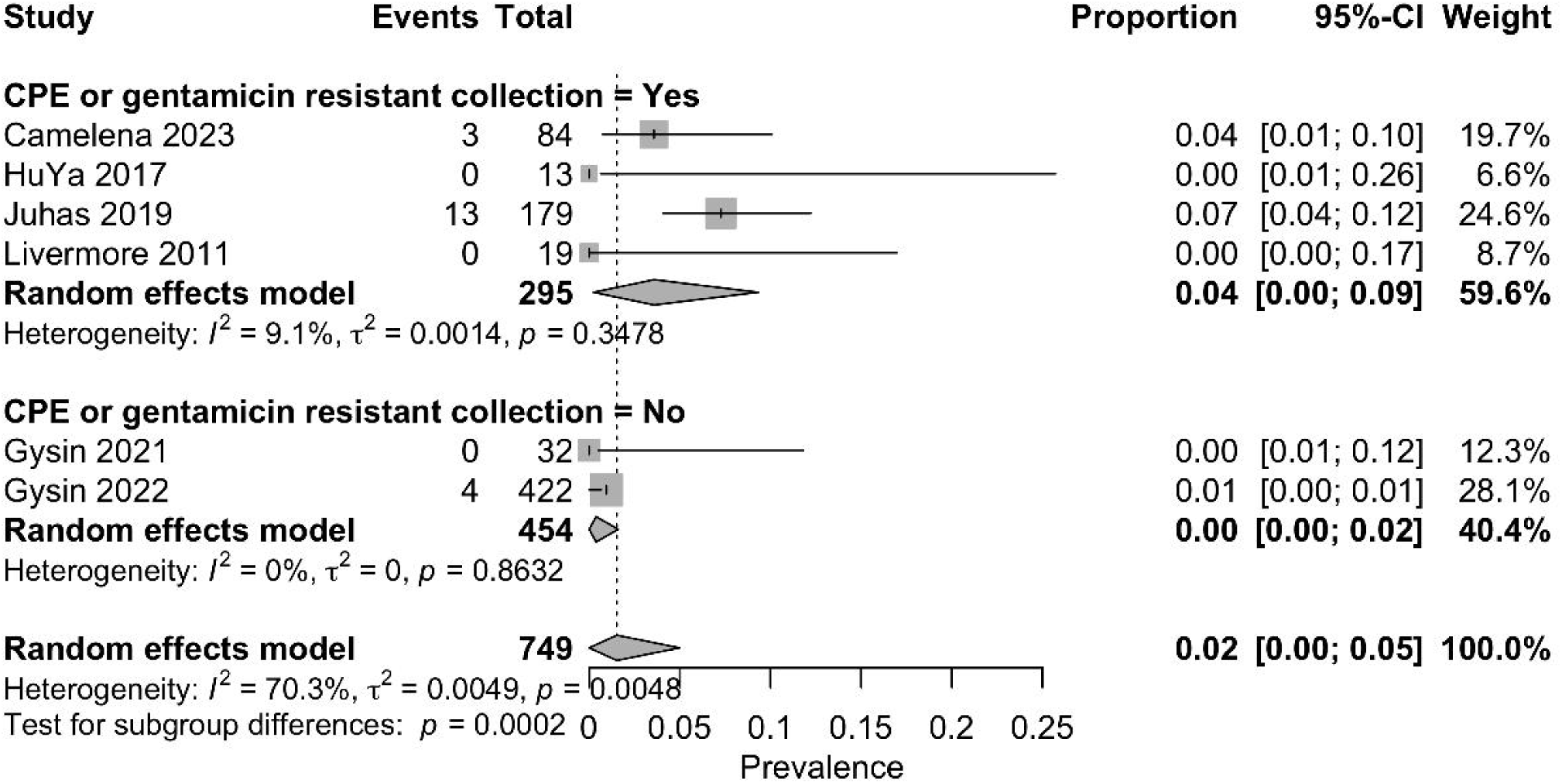
Meta-analysis presenting the pooled prevalence and 95% confidence intervals for apramycin resistance in *Enterobacter* spp (n=3 isolate collections)

**Figure 7:**
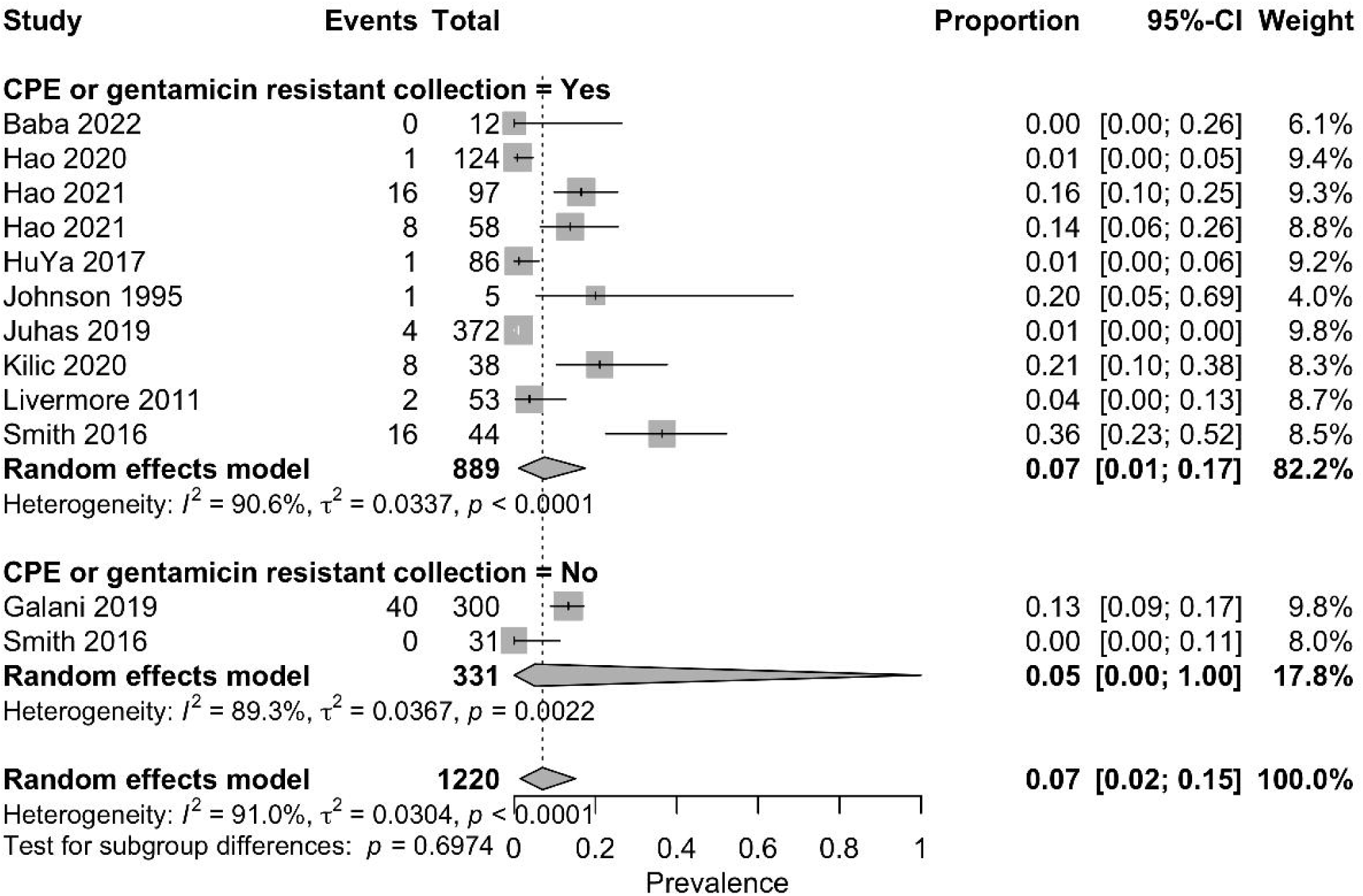
Meta-analysis presenting the pooled prevalence and 95% confidence intervals for apramycin resistance in *Klebsiella* spp (n=11 isolate collections)

**Figure 8:**
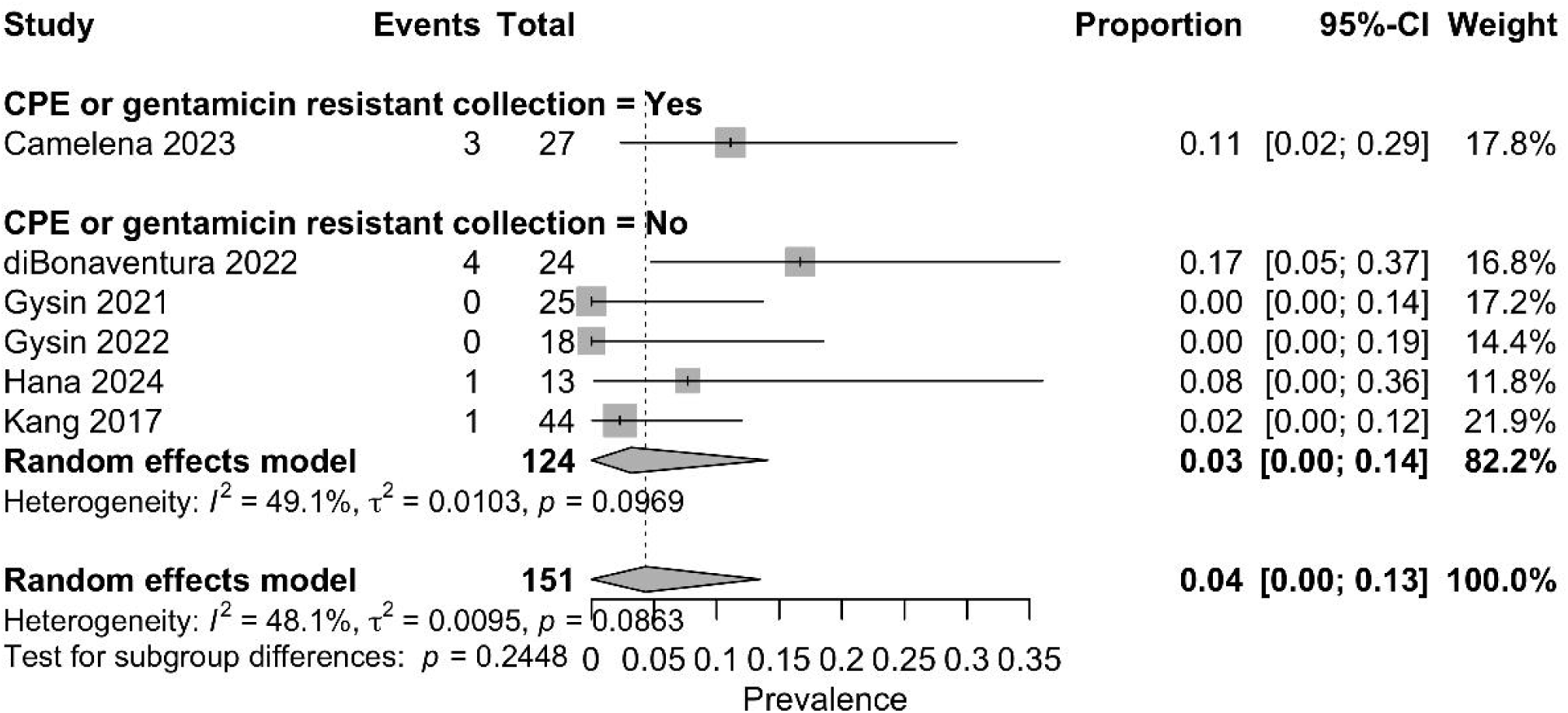
Meta-analysis presenting the pooled prevalence and 95% confidence intervals for apramycin resistance in *Pseudomonas* spp (n=6 isolate collections)

**Figure 9:**
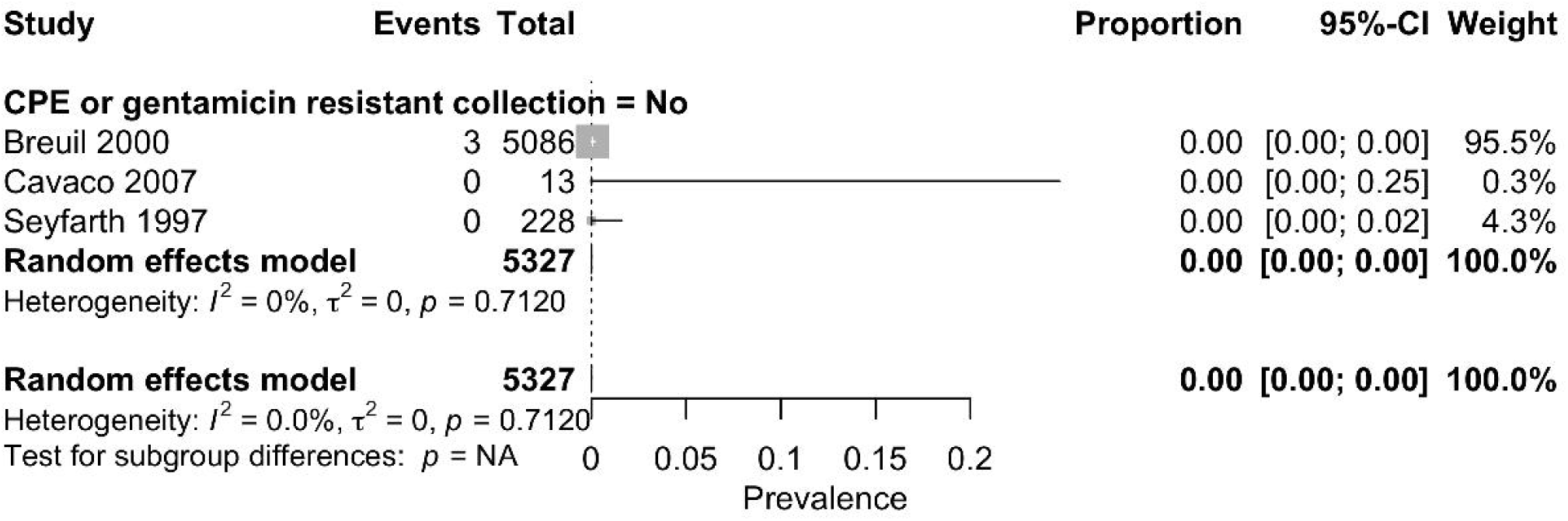
Meta-analysis presenting the pooled prevalence and 95% confidence intervals for apramycin resistance in *Salmonella* spp (n=3 isolate collections)

**Figure 10:**
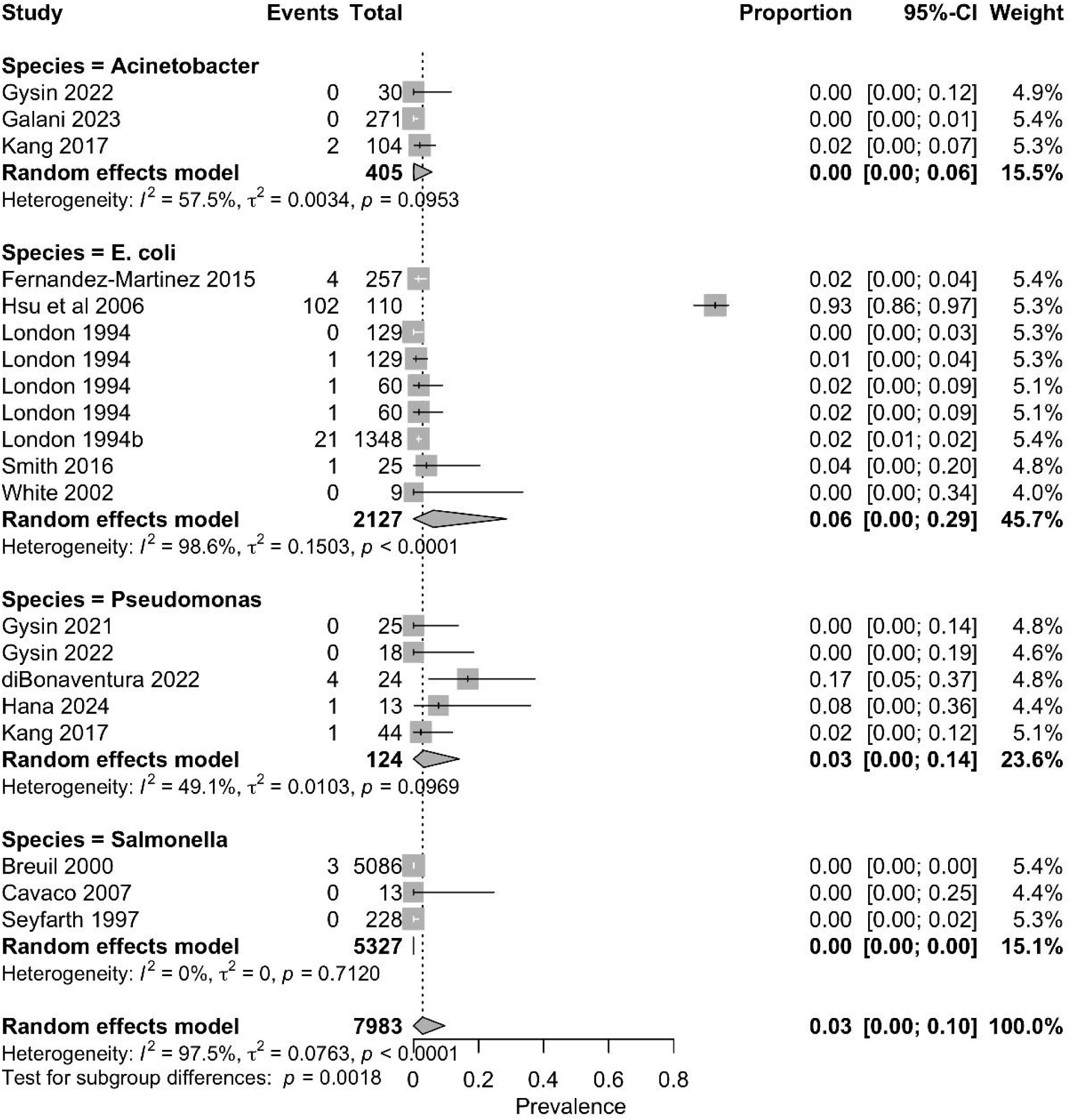
Meta-analysis presenting the pooled prevalence and 95% confidence intervals for apramycin resistance for 20 isolate datasets that were not selected on the basis of known carbapenem or gentamicin resistance (n=20)

Separate subgroup analysis was performed evaluating *E. coli* datasets known to be gentamicin resistant, carbapenem resistant, both or neither (Figure 11). A meta-regression of these 16 studies showed that the pooled prevalence of apramycin resistance in purposefully enrolled known gentamicin resistant isolates was significantly greater than those not purposefully selected as gentamycin resistant (*P*=0.03). There was no such association for carbapenem resistant isolate collections (*P*=0.63).

**Figure 11:**
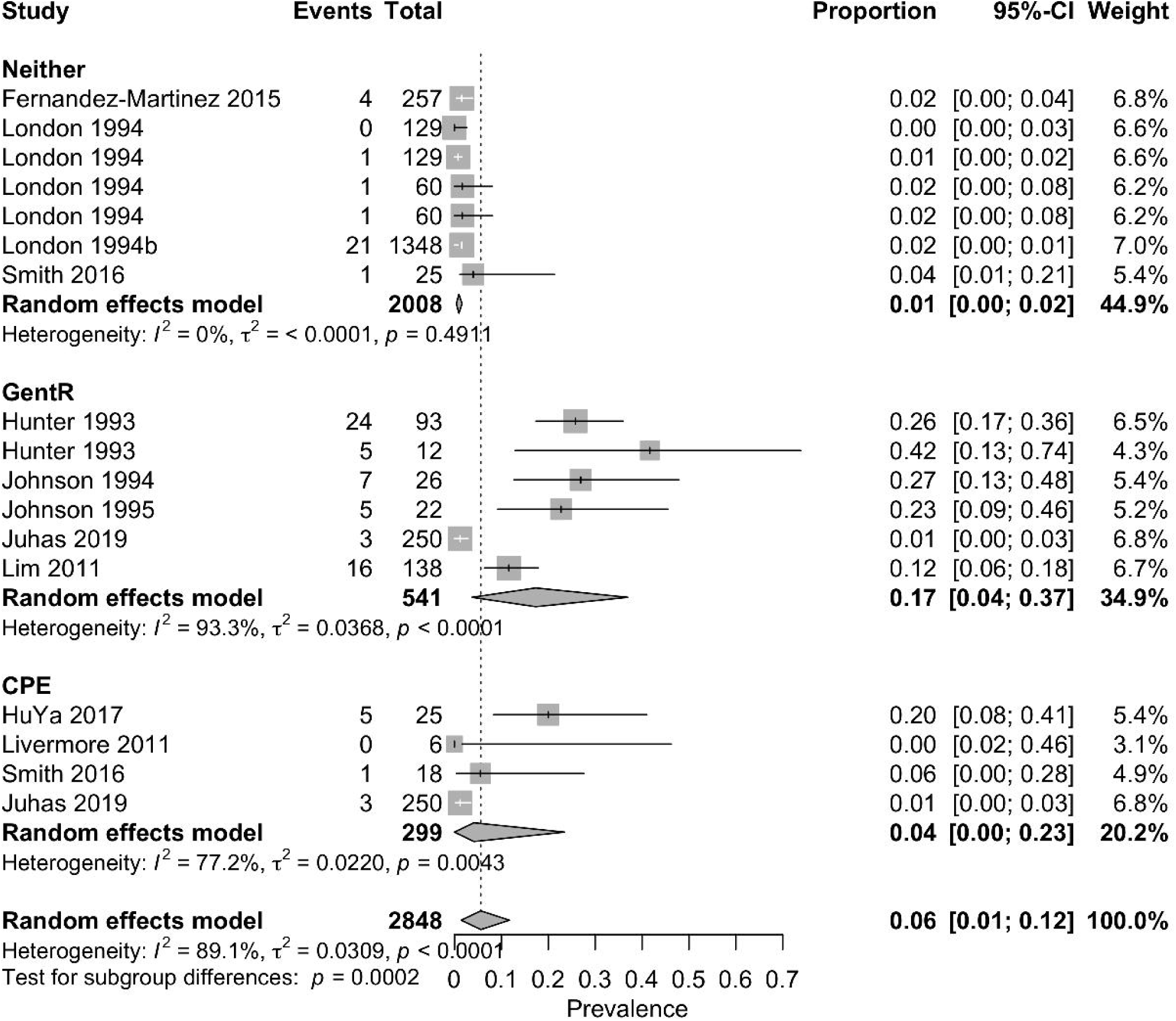
Meta-analysis presenting the pooled prevalence and 95% confidence intervals for apramycin resistance in *Escherichia coli* from isolate collections that were gentamicin resistant (GentR, n=5), carbapenem resistant (CPE, n=3), both carbapenem and gentamicin resistant (Both, n=1) or not specifically selected for gentamicin or carbapenem resistance (Neither, n=8). AMR: antimicrobial resist

*CoCoPop2: Are there differences in the prevalence of apramycin resistance in different bacterial species isolated from humans?*

This analysis was performed for bacterial species that were represented by 3 or more isolate collections that were not purposefully selected for resistance. Pooled estimates were determined for 20 datasets that reported resistance rates in *Acinetobacter* (n=3 study populations), *E. coli* (n=9), *Pseudomonas* (n=5) and *Salmonella* (n=3). A mixed effects meta-regression model was used and compared to the referent (*Acinetobacter*). There was no difference for *E. coli* (*P*=0.36), *Pseudomonas* (*P*=0.52) or *Salmonella* (*P*=0.90). This question was also explored using multivariable meta-regression, as is described below.

*CoCoPop-T: Has the prevalence of apramycin resistance in bacteria isolated from humans changed over time?*

Because of the significant impact of bacterial species and whether or not the isolate collection was known to be multidrug resistant, evaluation of the impact of time required study of individual bacterial species with isolate collections, and selected as CPE or gentamicin resistant, or not. There were inadequate numbers for any bacterial species/isolate collection type combinations, so this question could not be addressed at the bacterium level. The effect of year was evaluated using multivariable analysis, as described below.

### Multivariable analysis

Multivariable mixed-effects meta-regression was performed on the full dataset (excluding the outlier described above) and no effect of year (*P*=0.36), bacterial species (all *P*>0.19), geographic region (all *P*>0.13) or enrollment of known carbapenem-resistant isolates (*P*=0.44) was found. The only significant variable was datasets that used known gentamicin-resistant isolates (*P*=0.003).

When multivariable analysis was performed only on datasets not enrolled as gentamicin or carbapenem resistant, there was no effect of year (*P*=0.79), geographic region (all *P*>0.19) or bacterial species (all *P*<0.10).

## Discussion

Apramycin resistance was observed but was generally rare in bacteria isolated from humans. Pooled apramycin resistance prevalence estimates for all of the tested bacteria were low, typically <1% for isolate collections not selected on the basis of known gentamicin or carbapenem resistance. That resistance rate is consistent with a study that analyzed over 500,000 genomes of Gram-negative bacteria in the NCBI National Database of Antibiotic Resistant Organisms and identified *aac(3)-IV* in only 0.7% of isolates.^9^

The prevalence of resistance was higher in collections of known carbapenem-resistant and gentamicin-resistant strains but was still low. The highest pooled prevalence for this subpopulation was a rate of 14% in *E. coli*. When looking only at known gentamicin resistant *E. coli* isolate collections, the pooled prevalence of apramycin resistance was still only 17% (4-37%), with a prevalence of resistance of only 4% (0-23%) in carbapemase producing *E. coli*. The low apparent prevalence, even amongst multidrug resistant bacteria, highlights the unique structure of apramycin and its resistance to degradation by typical aminoglycoside resistance mechanisms, and correspondingly the potential value of repurposing apramycin for clinical use in humans.

Few associations were identified with apramycin resistance. This is probably in part because of the limited number of studies and heterogeneity in study populations and results. Lack of an identifiable effect of year is noteworthy. While the power to detect a difference may have been limited, low rates of resistance and lack of an apparent increase over time suggest that there is currently limited selection pressure towards acquisition and maintenance of apramycin resistance, from animal sources or otherwise.

While resistance rates were low, it is interesting to consider why there is resistance to a drug that is not used in humans. The predominant apramycin resistance gene, *aac(3)-IV* can be found in isolates from humans and animals,^10–13^ and whether or not resistance relates to veterinary use is unclear. This requires study if there may ultimately be dual use of this drug in human and veterinary medicine. Direct animal-human transmission of apramycin resistant bacteria has been reported on farms,^14^ and exposure to animal-origin apramycin resistant bacteria or apramycin resistance genes on mobile genetic elements through food, water or the environment could account for broader human community exposure. However, if there was significant resistance pressure from current and historical use of apramycin in livestock, it is reasonable to assume that resistance would be most common in bacteria that are more likely to be zoonotic in origin (e.g. *E. coli, Salmonella*) than those that are more human associated (e.g. *Klebsiella pneumoniae, Acinetobacter*). However, such a pattern was not noted here, with no difference in resistance between species. Indeed, apramycin resistance was rare in *E. coli* and *Salmonella* from sample collections not selected for multidrug resistance, despite use of apramycin in livestock for nearly 50 years. Results for *Salmonella* are particularly noteworthy as apramycin resistance was only identified in 3/5327 isolates. The number of studies involving *Salmonella* was small (n=3); however, the largest study was from France and reported a prevalence of 0.06% from over 5000 isolates. This involved samples collected between 1994-1997 in France,^15^ over a decade after apramycin use was first reported in livestock in that country.^16^ A recent systematic review of apramycin resistance in bacteria from livestock reported the pooled prevalence of apramycin resistance in *Salmonella* was 4% in healthy livestock and 7% in diseased livestock (Weese et al, unpublished data). While still low, this is in contrast with the rarity of resistance identified here. Data from this study cannot indicate whether or not there is any influence of veterinary use on apramycin resistance in human isolates, but they show that resistance is currently rare, with no apparent increased risk in bacteria that would more likely be of zoonotic origin or increasing rates over time.

It is possible that aminoglycoside use in humans could select for apramycin resistance independent of any animal link. If *aac(3)*-IV is present in human commensals and pathogens, there would be some degree of selection pressure from use of other aminoglycosides in humans. However, aminoglycoside use in humans would be more likely to select for common aminoglycoside resistance genes such as *aac(3)-II, aac(6*′*)-I, ant(3*″*)-I, aph(3*′*)-II,* and *ant(2*″*)-I,* not *aac(3)-IV,* which do not confer apramycin resistance. In the current situation where *aac(3)-IV* is rare in isolates from humans, it is reasonable to suspect that there would be limited selection pressure for apramycin resistance from use of non-apramycin aminoglycosides. If apramycin resistance genes become more common, from human or animal influences, the selection pressure from any aminoglycoside use could increase.

In addition to the small number of studies of different bacteria, other limitations and knowledge gaps must be considered. There are no established interpretive breakpoints for apramycin resistance in bacteria from humans. Some studies reported their breakpoints, some used epidemiological cutoff (ECOFF) interpretation and some did not report their criteria. When breakpoints were reported, these differed somewhat between studies. Whether this would have impacted results is unclear as minimum inhibitory concentration (MIC) data were not typically reported, precluding re-interpretation using a standard breakpoint. Establishment of breakpoints is needed for future clinical use and surveillance.

Studies that did not purposefully enrol resistant isolates still may have been biased toward resistance, such as studies from patients with cystic fibrosis, hospital-associated infections or multidrug resistant (but not known carbapenem or gentamicin resistant) isolate collections. This would be assumed to bias results towards over-estimation of resistance in the lower risk population, so actual resistance in the community may be even lower than the low rates that were observed here. Regional differences in resistance may also be present but investigation was limited here because of the small number of studies from different regions.

Apramycin resistance was identified in bacteria of human origin but was rare, supporting the potential value of re-purposing this drug for use in humans. Despite nearly 50 years of use in animals, resistance rates in zoonotic pathogens were very low, suggesting that there is limited spillover of resistance from veterinary and agricultural use of apramycin. However, further monitoring of apramycin resistance would be warranted should it become authorized for use in humans, to detect changes in resistance and potential risk factors for resistance development and dissemination. Further surveillance is needed to better understand apramycin resistance in different bacterial and human populations and factors that influence resistance, as well as the relationship between apramycin use and resistance in animals, and resistance in humans.

## Data Availability

All data produced in the present study are available upon reasonable request to the authors

